# A longitudinal study of age-related traits and cognitive function: The Collaborative Amish Aging and Memory Project (CAAMP)

**DOI:** 10.64898/2026.09.17.26363351

**Authors:** Dana Z. Jian, Renee A. Laux, Yeunjoo E. Song, Audrey Lynn, Kristy Miskimen, Alex Gulyayev, Sarada L. Fuzzell, Sherri D. Hochstetler, Dawn Miller, Penelope Miron, Laura J. Caywood, Jason E. Clouse, Sharlene D. Herington, Michael B. Prough, Larry D. Adams, Yining Liu, Noel C. Moore, Daniel A. Dorfsman, Paula Ogrocki, Alan J. Lerner, Jeffery M. Vance, Michael L. Cuccaro, Margaret A. Pericak-Vance, William K. Scott, Jonathan L. Haines

## Abstract

**INTRODUCTION:** The Collaborative Amish Aging and Memory Project (CAAMP) is an ongoing longitudinal study spanning multiple sites, focused on cognitive function and age-related traits in Midwestern Amish (Ohio and Indiana) communities. The primary traits of interest include Alzheimer Disease and related dementias (ADRD), successful aging (SA), and other age-related conditions.

**METHODS:** CAAMP integrates clinical and cognitive assessments with family history, genealogical, genomic, biospecimen, and biomarker data collected across Amish communities in Ohio and Indiana. Cognitive diagnoses are assigned through consensus adjudication using clinical, cognitive, functional, and informant data.

**RESULTS:** To date, CAAMP has enrolled 3,978 participants (mean age 78.6 (SD = 7.9), 59.1% female, and 83% with an education level at the eighth grade), of whom 2,928 have consensus-adjudicated cognitive status. At the most recent assessment, 66.6% were cognitively unimpaired, 9.9% had mild cognitive impairment, 8.9% had Alzheimer disease, and 4.5% had cognitive impairment, but not Alzheimer disease. The remaining 1,050 did not have detailed cognitive evaluations at their enrollment. Longitudinal cognitive assessments and multiple biospecimen sample types are available for subsets of participants. CAAMP further includes extensive multigenerational pedigree information, genome-wide genotype and whole-genome sequencing data, and plasma biomarker measurements.

**DISCUSSION:** CAAMP illuminates the genetic underpinnings of cognition and late life diseases, particularly Alzheimer Disease, by leveraging the special characteristics of a founder population. Continued study of this prospective cohort provides opportunities to identify the genetic and biological factors driving cognitive outcomes alongside other age-related conditions, advancing our understanding of both disease risk and protection.

## 1. Introduction

### 1.1 Advantages of Founder Populations

Genetic and genetic epidemiology studies aim to identify DNA variation associated with disease risk and protection. Family-based studies are particularly well suited for the discovery of rare or moderately frequent variants with larger effect sizes, and for disentangling genetic effects from shared environmental influences.^1^

The Amish represent a unique founder population especially well-suited for family-based research and that offers important opportunities for studying the genetic architecture of age-related diseases. Detailed multigenerational genealogical records enable efficient identification and study of extended pedigrees. As a founder population, the Amish are characterized by reduced genetic heterogeneity, increased relatedness, extended linkage disequilibrium, and enrichment of rare or moderately frequent variation. These features enhance power to detect recessive, non-additive, and protective genetic effects that may be difficult to identify in more heterogeneous populations or population-based cohorts of unrelated individuals.

Amish communities offer distinct research advantages, including large multigenerational families, stable residency patterns, and relatively consistent environmental exposures. These factors facilitate family ascertainment and longitudinal follow-up. Together, this combination of genealogical depth, genetic structure, and community stability provides a strong foundation and opportunity for family-based and longitudinal genetic studies, particularly for complex, age-related conditions.

### 1.2 History, Religious Context, and Sociocultural Norms

As a founder population, the Amish are culturally, religiously, and genealogically distinct, living primarily in rural regions of the United States. They descend from German and Swiss Anabaptist Christians who immigrated in two major waves between 1727-1770 and 1815-1860.^2^ As the population expanded, large Midwestern settlements developed in Ohio, Indiana, and Pennsylvania.^3^ These settlements reflect distinct genealogical origins: communities in Holmes and surrounding Counties, Ohio, and Elkhart/LaGrange and surrounding Counties, Indiana, primarily trace their ancestry to earlier immigrants from the German Palatinate, whereas the Adams County, Indiana community traces to later Swiss immigrants.^2,4^

Amish life is centered on religious and distinct sociocultural norms transferred across generations. Although practices vary across orders and church districts, each governed by its own *Ordnung* (the unwritten community rules guiding religious practice, technology use, and daily life), Amish communities share core Anabaptist commitments emphasizing faith, reconciliation, humility and collective well-being. Strong kinship and congregational networks, distinctive dress, and use of German-derived vernaculars alongside English, further characterize Amish life. Although historically agrarian, occupations have diversified to include farming, family-owned business, skilled trades, manufacturing and employment in larger enterprises. The Amish typically operate their own schools, with most individuals discontinuing formal education after the eighth grade. However, many individuals acquire substantial informal education through apprenticeships, occupational training, and experiential learning associated with their trades or businesses.

Religious and community values also influence health-related decision-making. While modern medicine is generally accepted, particularly for traumatic or obstetrical care, many individuals also draw upon traditional and cultural health knowledge, folk remedies, spiritual practices, family care and support, community or church-based aid, specialized Amish clinics, or alternative health professionals when available.^5^ Barriers to care vary due to rural residence, transportation limitations, technology restrictions, cost, and personal or cultural preferences.^5^ Despite these challenges, Amish communities often demonstrate comparable or longer life expectancy than the general U.S. population^6^, potentially reflecting lifestyle and social factors unique to Amish culture.

Altogether, this distinct religious, cultural, and historical background provides important context for understanding health, research participation, and interpretation of disease patterns within Amish populations.

Alongside this sociocultural context, the genetic, genealogical, and familial characteristics of the Midwestern Amish provide unique opportunities for research on aging and age-related diseases. These scientific advantages have served as the foundation for supporting more than three decades of research that have evolved into the Collaborative Amish Aging and Memory Project (CAAMP), an ongoing, multisite study of Alzheimer disease and related dementias (ADRD), cognitive preservation, successful aging, and other age-related conditions. This paper describes the development of CAAMP and its study protocols, characterizes the cohort and research resources, summarizes major scientific contributions, and discusses opportunities for continued investigation of genetic and non-genetic factors related to cognitive function and aging.

## 2. Methods

### 2.1 Engaging Amish communities: The importance of cultural responsiveness

Amish communities are not uniform and generalizations about Amish beliefs, behaviors, or practices, like those made about any population, are neither accurate nor appropriate. Substantial variation exists across individuals, families, church districts, affiliations, and generations. Rather than treating Amish identity as an all-encompassing category, it is more accurate to view it as one element among many, including family networks, local community norms, occupational contexts, and religious and migration histories.

Trust between Amish communities and CAAMP’s research team has been established and sustained through long-term engagement and intentional cultural responsiveness. In the earliest phases of CAAMP, the research team engaged directly with Amish bishops and other respected community members to understand local norms, religious values, and community expectations before initiating enrollment. These discussions led to an approach that prioritizes individual decision making, such that participants are engaged directly, identified using published Amish community directories and approached at their homes. Some constituent studies targeted families with specific phenotypes or known variants, while others approached eligible individuals without regard to family history.

These conversations are revisited whenever study protocols are modified or new measures are introduced, particularly for sensitive topics requiring careful cultural framing. Community input ensures that study procedures and assessments are conducted in a manner that is respectful, understandable, and acceptable within specific contexts. For example, the photo of the witch was removed from the Multilingual Naming Test (MINT) as a culturally appropriate adaptation. In addition, research staff conduct study interactions in ways that reflect local cultural norms, including attention to dress, communication style, and behavior during home visits and community-based assessments. These practices support clear communication, reinforce respect for participant autonomy, and reflect an ethic of cultural humility rather than presumption. This approach has been central to sustaining participation and collaboration for more than three decades.

Across CAAMP’s studies, participants are not routinely provided monetary compensation. Participation is most often motivated by trust in the research team, perceived benefit to family, community, or humankind, and a willingness to contribute to research rather than any financial incentive.

### 2.2 Development of CAAMP

From both scientific and population health perspectives, research involving Amish populations has a long history and the Amish are recognized as an informative population. Genetic studies on the Amish and other Anabaptist populations began in the 1960s^7^, with the identification and characterization of rare Mendelian disorders, establishing these communities as informative populations for medical genetics.^8^ Subsequent research expanded to complex and more common diseases.^8^ Additionally, the communities’ openness to participation, combined with the critical role of cultural liaisons, have enabled the creation of extensive pedigrees, family histories and genealogical databases that are valuable resources today.^9,10^

The focus on Alzheimer disease (AD) and cognitive aging research in the Midwestern Amish began with a population-based epidemiologic survey conducted between 1991 and 1993 across four Amish settlements around Northern Indiana (Elkhart, LaGrange, Adams, and St. Joseph Counties).^11^ In this survey, 632 eligible adults aged ≥65 years were identified, of whom 516 completed the Mini-Mental State Examination (MMSE).^11,12^ The prevalence of probable dementia was estimated at 6.4%, substantially lower than that observed in similarly aged U.S. populations.^11^

Concurrently, advances in Alzheimer genetics, including the 1993 landmark discovery of the apolipoprotein E (*APOE*) ε4 allele as a strong genetic risk factor for sporadic late-onset AD (LOAD)^13^, followed by the identification of the *APOE* ε2 allele as protective^14^, prompted early genetic investigations of AD in the Midwestern Amish. In a 1996 initial family-based study of an extended Amish pedigree with multiple LOAD cases and community controls, we observed a low *APOE* ε4 allele frequency (∼ 4%), with all affected individuals carrying the *APOE* ε3/ε3 genotype.^15^ These findings suggested that the lower frequency of *APOE* ε4 may partially contribute to the reduced prevalence of dementia in the Midwestern Amish, yet at the same time suggesting that other genetic variation might be important, underscoring the scientific value of studying AD in this founder population.

Encouraged by these initial findings, our AD research efforts expanded to include more systematic ascertainment of affected and unaffected individuals across Amish communities in Indiana and Ohio. This expansion incorporated standardized cognitive assessments, structured clinical interviews, and family-based recruitment strategies. Early pilot funding and subsequent federal support in the early 2000s enabled broader investigation of cognitive aging and successful aging, along with coordinated data collection across multiple sites using population-based and family-based ascertainment designs.

Building on this foundation, the research evolved into a long-standing, multi-decade, multi-site effort. This work supported investigations of successful aging, cognitive trajectories, dementia risk, and related age-associated conditions, including Parkinson’s disease (PD), age-related macular degeneration (AMD) and glaucoma. Sustained funding from NIH and private foundations facilitated continued recruitment, protocol refinement, and expansion of longitudinal follow-up.

Collectively, these efforts represent a continuous research lineage now consolidated as the Collaborative Amish Aging and Memory Project (CAAMP), an ongoing multi-site, longitudinal study designed to investigate cognitive decline, dementia risk, successful aging, and other age-related traits within Midwestern Amish populations (Figure 1).

**Figure 1.**
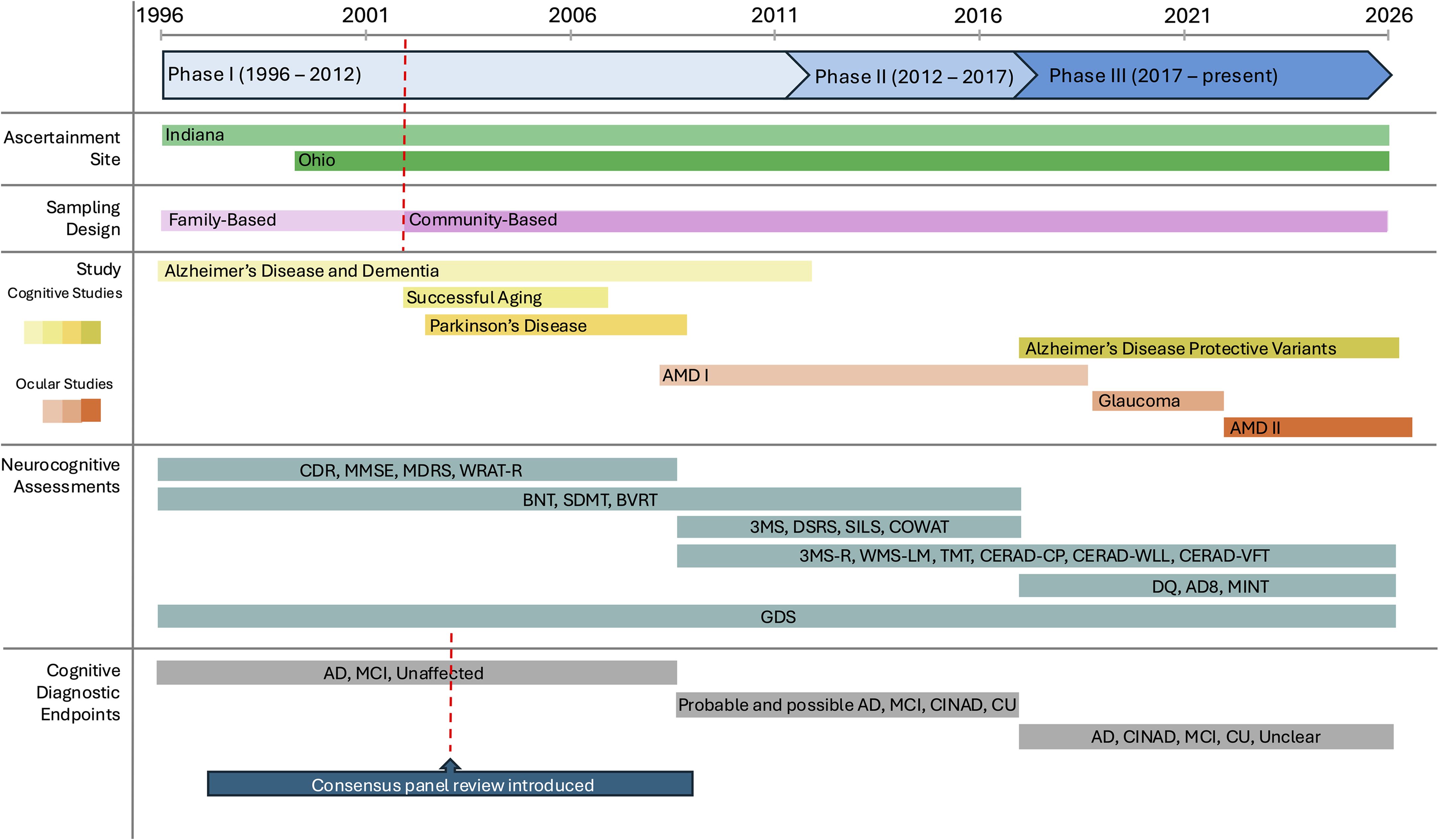
CAAMP Study Timeline, 1996 – 2026 (current). CAAMP currently spans three phases: Phase I (1996-2012), Phase II (2012-2017) and Phase III (2017 – present) with ascertainment in Indiana and Ohio. Recruitment transitioned from family-based to primarily community-based design around 2006-2007. The figure summarizes cognitive and ocular studies, neurocognitive assessment batteries and diagnostic endpoints across the study period. Consensus-based adjudication began in 2003. Bars indicate periods of activity or use, and red dashed lines indicate major study transitions. Funding periods are provided in Supplementary Table 2. Abbreviations: CDR = Clinical Dementia Rating Scale; MMSE = Mini-Mental State Examination; MDRS = Mattis Dementia Rating Scale; WRAT-R = Wide Range Achievement Test-Revised; BNT = Boston Naming Test; SDMT = Symbol Digit Modalities Test; BVRT = Benton Visual Retention Test; 3MS = Modified Mini-Mental State Examination; DSRS = Dementia Severity Rating Scale; SILS = Shipley Institute of Living Scale; COWAT = Controlled Oral Word Association Test; 3MS-R = Modified Mini-Mental State Examination-Revised; WMS-LM = Wechsler Memory Scale Logical Memory; TMT = Trail Making Test A and B; CERAD-CP = CERAD Constructional Praxis; CERAD-WLL = CERAD Word List Learning; CERAD-VFT = Consortium to Establish a Registry for Alzheimer’s Disease Verbal Fluency Test; DQ = Dementia Questionnaire; AD8 = AD8 Dementia Screen; MINT = Multilingual Naming Test; GDS = Geriatric Depression Scale. AD = Alzheimer’s disease dementia; MCI = Mild cognitive impairment; CINAD = cognitively impaired not Alzheimer disease; CU = cognitively unimpaired.

### 2.3 Clinical Protocol for Cognitive Measures

The core study entry requirements and inclusion criteria have remained consistent throughout the 3+ decades of CAAMP: all participants are required to be genetically related to these Amish communities (with almost all being active members), to express willingness to participate, provide a biological sample, and be 60 years of age or older (with exceptions for younger family members, per specific protocol). All subjects have provided informed consent, either directly or via a proxy, and all studies have been approved by the Institutional Review Boards of the appropriate institutions.

Upon enrollment, all participants complete a standardized set of interviews, clinical assessments, and surveys. While the specific measures have evolved and expanded over time^16,17^, core domains have remained consistent. Assessments are typically conducted in participants’ homes or, when preferred, at a community-based assessment center. With participant permission, family members, most often spouses or adult children, are commonly present and frequently serve as informants.

Across study phases, collected data have included sociodemographic characteristics, medical and family history, neurocognitive abilities, functional status, and neuropsychiatric symptoms. All available clinical and informant-based information has been integrated to adjudicate diagnostic endpoints, which have evolved according to study phase and prevailing diagnostic standards.

#### 2.3.1 Evolution of Clinical Assessment Methods

Clinical assessment methods and protocols have varied across phases of CAAMP (Table 1) and, as appropriate, have been adjusted for the educational characteristics of the Amish population (details in Supplemental Material 1 and Supplementary Table 1). Additional details and references for functional, physical, behavioral, and other non-cognitive measures are provided in Supplemental Material 1.

**Table 1.**
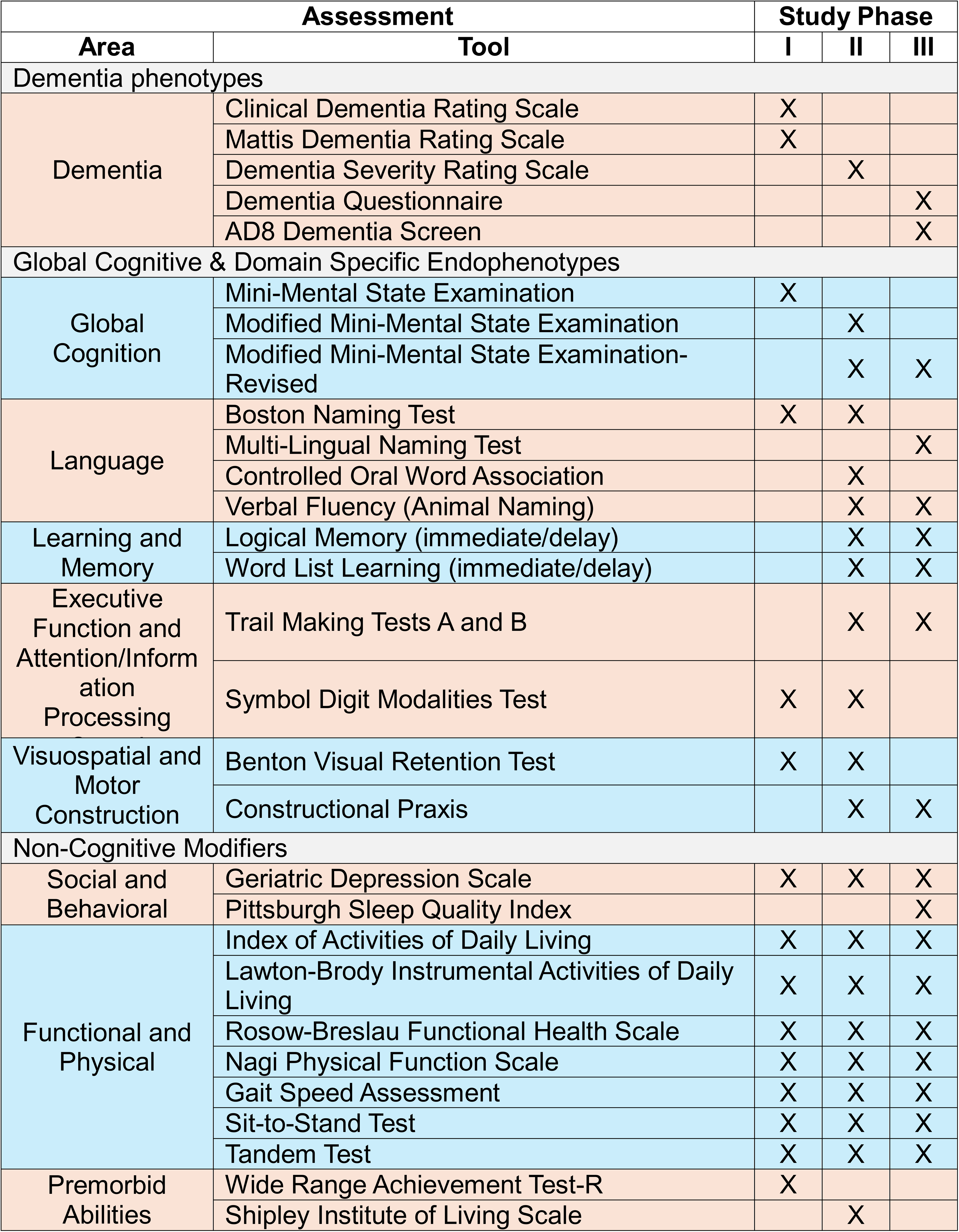
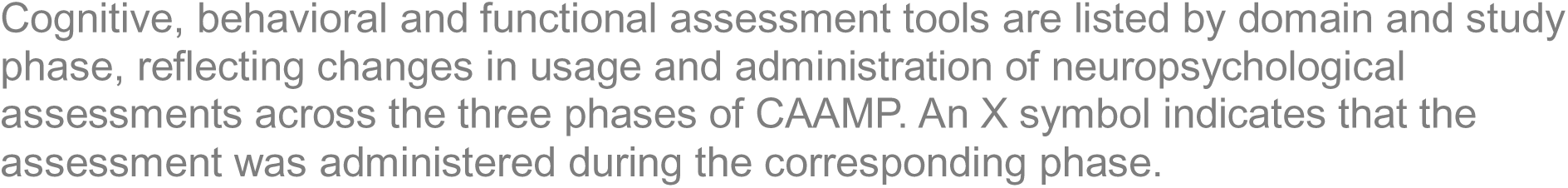
Clinical Assessments by Study Phase.

**Phase I (1996-2012):** Clinical measurement was used to distinguish cognitively unimpaired individuals from those with cognitive impairment.^16^ Domains of interest included family and medical history, assessments of daily functioning, and the Geriatric Depression Scale (GDS), a self-report measure of depressive symptoms.^18^ Functional and physical assessments included measures of activities of daily living, mobility and physical function, and physical performance; these assessments were retained in subsequent study phases (Table 1; see Supplemental Material 1 for additional details and instrument references).

Cognitive screening was performed using the Mini-Mental State Examination (MMSE^12^), a 30-item global cognitive screening instrument. Participants with MMSE scores >27 were considered as cognitively unimpaired, while those with scores <24 were considered as cognitively impaired consistent with dementia. Participants with intermediate scores (MMSE 24–26) or evidence of impairment were administered an expanded assessment battery, which included the Mattis Dementia Rating Scale (MDRS^19^) the Boston Naming Test (BNT^20^), the Wide Range Achievement Test (WRAT-R^21^) and additional clinical information related to memory function. This tiered approach aligned with standard epidemiologic practices (brief, broad screening test followed up with more detailed, specific testing) and produced well-defined diagnostic phenotypes or endpoints for subsequent analyses.

##### Phase II (2012-2017)

Screening procedures were updated with the replacement of the MMSE by the Modified Mini-Mental State Examination (3MS^22^), and later the revised version (3MS-R^23^). The tiered assessment approach was retained: individuals with 3MS-R scores >87 was classified as cognitively unimpaired, while those scoring <87 underwent further evaluation using an expanded neuropsychological battery.

This battery included the Consortium to Establish a Registry for Alzheimer Disease (CERAD) neuropsychological measures^24^, supplemented by memory measures from the Wechsler Memory Scale-Logical Memory subtest (WMS^25^), the Benton Visual Retention Test (BVRT^26^), Trail Making Tests A and B (TMT^27^), and the Controlled Oral Word Association Test (COWAT^28^). The Shipley Institute of Living Scale (SILS^29^) was included to assess premorbid intellectual functioning. As in earlier phases, the GDS was used to assess depressive symptoms. Dementia severity was quantified using the Dementia Severity Rating Scale (DSRS^30^).

##### Phase III (2017-Present)

The most recent phase of CAAMP has retained several previously used measures, with adaptations and the addition of new instruments. This phase initially emphasized enrollment of cognitively unimpaired individuals, reflecting a primary focus on identifying genetic variants associated with protection against dementia. The 3MS-R continues to serve as the primary screening tool, and CERAD measures, including Word List Learning (CERAD-WLL) and Constructional Praxis (CERAD-CP), have been maintained^24^.

Several measures were updated in this phase. The COWAT was replaced by the Semantic Fluency (animal naming) task, and the BNT was replaced by the Multilingual Naming Test (MINT^31^). In addition, for participants unable to complete the full neuropsychological battery, the Dementia Questionnaire (DQ^32^), a semi-structured, informant-based interview assessing cognitive and behavioral changes consistent with dementia, was administered to a family member along with the AD8 Dementia Screening Interview (AD8^33^), an informant-completed measure of dementia-related functional change. Assessment of depressive symptoms using the GDS and functional and physical status measures has been consistently maintained.

#### 2.3.2 Diagnostic Endpoints and Adjudication Procedures

Diagnostic classification adhered to diagnostic standards appropriate for each study period, including McKhann et al. 1984^34^, McKhann et al. 2011^35^, Albert et al. 2011^36^, and Jack et al. 2018.^37^

##### Phase I (1996-2012)

Based on available clinical data, participants were classified as dementia, mild cognitive impairment (MCI/unclear), or unaffected. Individuals classified as dementia were further classified as possible or probable AD according to the NINCDS-ADRDA criteria for Alzheimer disease^34^. These classifications were based on MMSE scores and cognitive testing for select cases. Research diagnostic assignments were assigned by clinicians who had conducted the assessments until 2003 at which time a consensus panel consisting of a neuropsychologist and nurse practitioner, both with geriatric expertise, reviewed all cases.

##### Phase II (2012-2017)

Diagnostic determinations shifted to consensus case conferences with detailed case reports prepared by study coordinators and reviewed by an expanded consensus panel consisting of a physician assistant, clinical nurse specialist, and two neuropsychologists. Following group discussion, consensus impressions were assigned, and age at onset was estimated for participants classified with dementia. Research diagnostic categories included probable Alzheimer disease, possible Alzheimer disease, MCI, cognitively impaired not Alzheimer disease (including vascular dementias), and cognitively unimpaired.

##### Phase III (2017-Present)

Case reports are reviewed by a panel composed of a clinical psychologist, a neuropsychologist, and two neurologists. Case reports are structured and include information from all relevant domains including sociodemographic information, family and medical history, functional information, and cognitive test results. Current research diagnostic categories include cognitively unimpaired (CU), mild cognitive impairment (MCI), Alzheimer disease dementia (AD), cognitively impaired not Alzheimer disease (CINAD), and unclear. For broader analyses, a cognitively impaired phenotype (CI) can be defined by combining individuals classified as MCI, AD or CINAD.

###### Consistency of Diagnostic Endpoints

A strength of the CAAMP clinical enterprise is the use of standard measures across all study phases. These measures along with clinical history and examiner impressions have provided a solid foundation for adjudication processing. Since a substantial number of participants have multiple visit data, the consensus panel has had the opportunity to review these prior visits and confirm the assigned endpoints.

#### 2.3.3 Collection of Biological Samples

While CAAMP’s study protocols have evolved over time, it has been standard practice to obtain a biological sample from all Amish participants. Blood samples have always been preferred; however, saliva, buccal swabs, or filter cards have been collected for a small number of individuals when blood collection was not feasible. Cell lines (lymphoblastoid cell lines (LCL) or induced pluripotent stem cells (iPSC)) have been generated on a small number of participants with notable traits (e.g. exceptional memory, *APOE* ε4/ε4 genotype, age 90+).

For cognitive studies, blood samples are typically obtained in temporal proximity to the cognitive assessment. CU subjects who are seen for follow-up visits provide an additional blood sample at each subsequent visit. For ocular studies, blood samples are collected on the day of the initial eye examination, but not on follow-up. In phase I, blood draws were performed only for participants with suspected cognitive impairment. The protocol was later modified to include sample collection from all participants, including those previously enrolled as CU. Additional details regarding sample collection are provided in Supplemental Material 2.

## 3. Results

### 3.1 Demographics

As of December 31, 2025, CAAMP has 3,978 participants, with approximately 48.5% of participants from the Ohio (OH) site; 59.1% are female and 83% have 8 years of formal education. Ohio and Indiana (IN) participants had a mean age of 77.67 (SD = 7.6) and 80.1 (SD = 8.2) years at their last exam, respectively; at first enrollment, the corresponding mean ages were 74.82 (SD = 6.67) and 77.49 (SD = 8.15) years. The distribution of age and education differed by site (*p < 0.001)* (Table 2). Among CAAMP participants with available *APOE* genotype information, 636 (24.8%) individuals were *APOE* ε4 carriers, while 1,929 individuals were non *APOE* ε4 carriers (Table 2). The distribution of *APOE* genotype differed by site (*p = 0.001*).

**Table 2.**
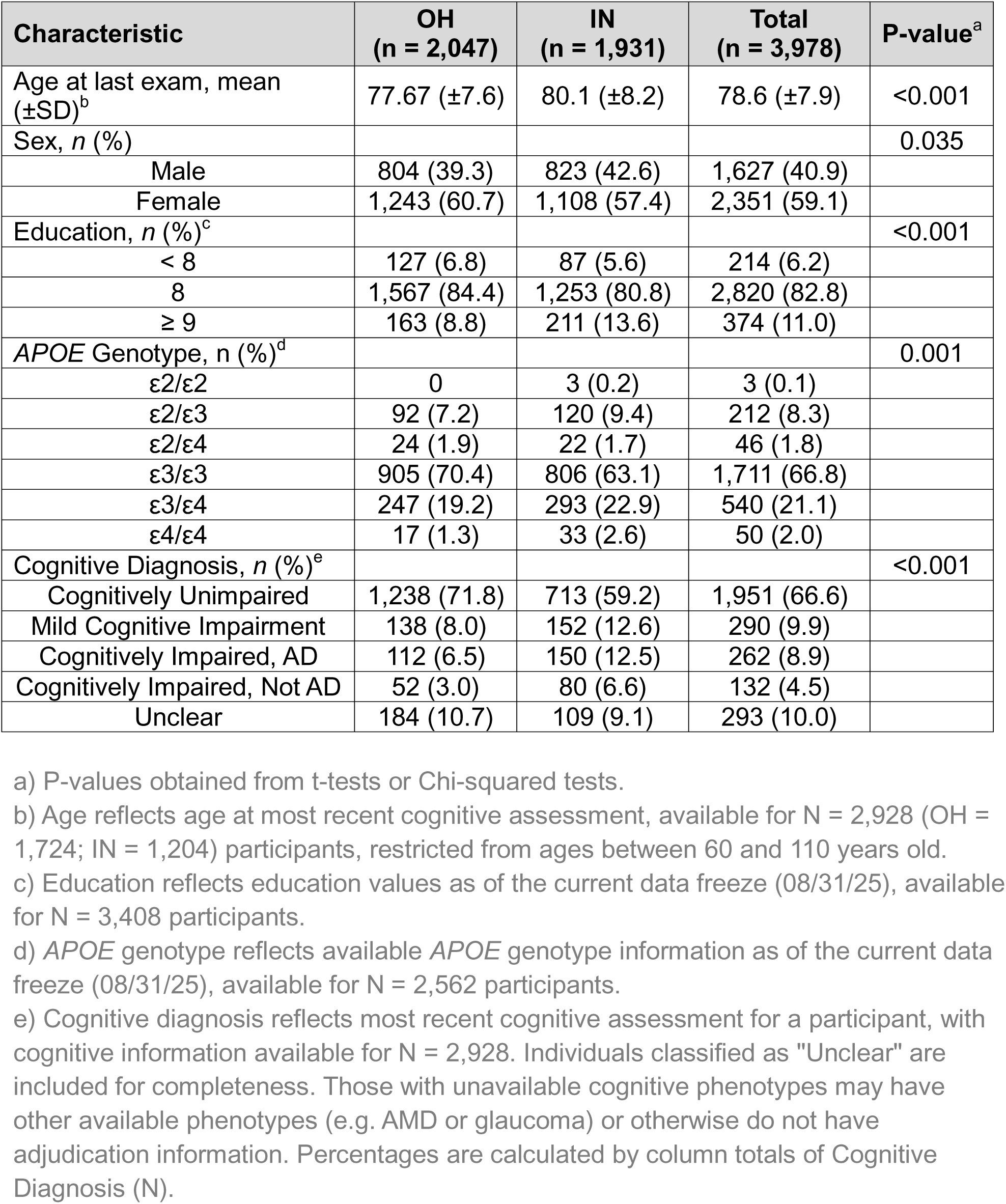
Descriptive Characteristics of the Mid-Western Amish.

For participants with available cognitive phenotype information (N = 2,928), diagnosis was based on most recent assessment. Of these, 66.6% are CU, 9.9% are MCI, 8.9% are AD. A smaller proportion, 4.5%, were classified as CINAD (non-AD etiologies). The distribution of cognitive phenotype differed by site (*p = 0.001*). To determine whether observed site differences reflected the higher prevalence of AD in Indiana, we fit regression models adjusting for AD diagnosis. After adjustment, Indiana participants remained significantly older (β = 1.95 years, *p < 0.001*) and had higher odds of carrying at least one *APOE* ε4 allele (OR = 1.45, p < 0.001) compared to Ohio participants, whereas the sex distribution did not differ by site (*p = 0.15*). Longitudinal cognitive trajectories are available for participants for up to four cognitive assessments. Among participants with repeated assessments, median follow-up time between consecutive examinations is 8.9, 2.8, 2.2, and 2.2 years, respectively (Figure 2). Previous evaluations of follow-up in CAAMP demonstrated an overall retention rate of 68.97%. Of those not retained, attrition is driven primarily by death (49.42%) or reaching study endpoints (40.6%), resulting in minimal loss to follow-up due to participant withdrawal or refusal (9.98%).

**Figure 2.**
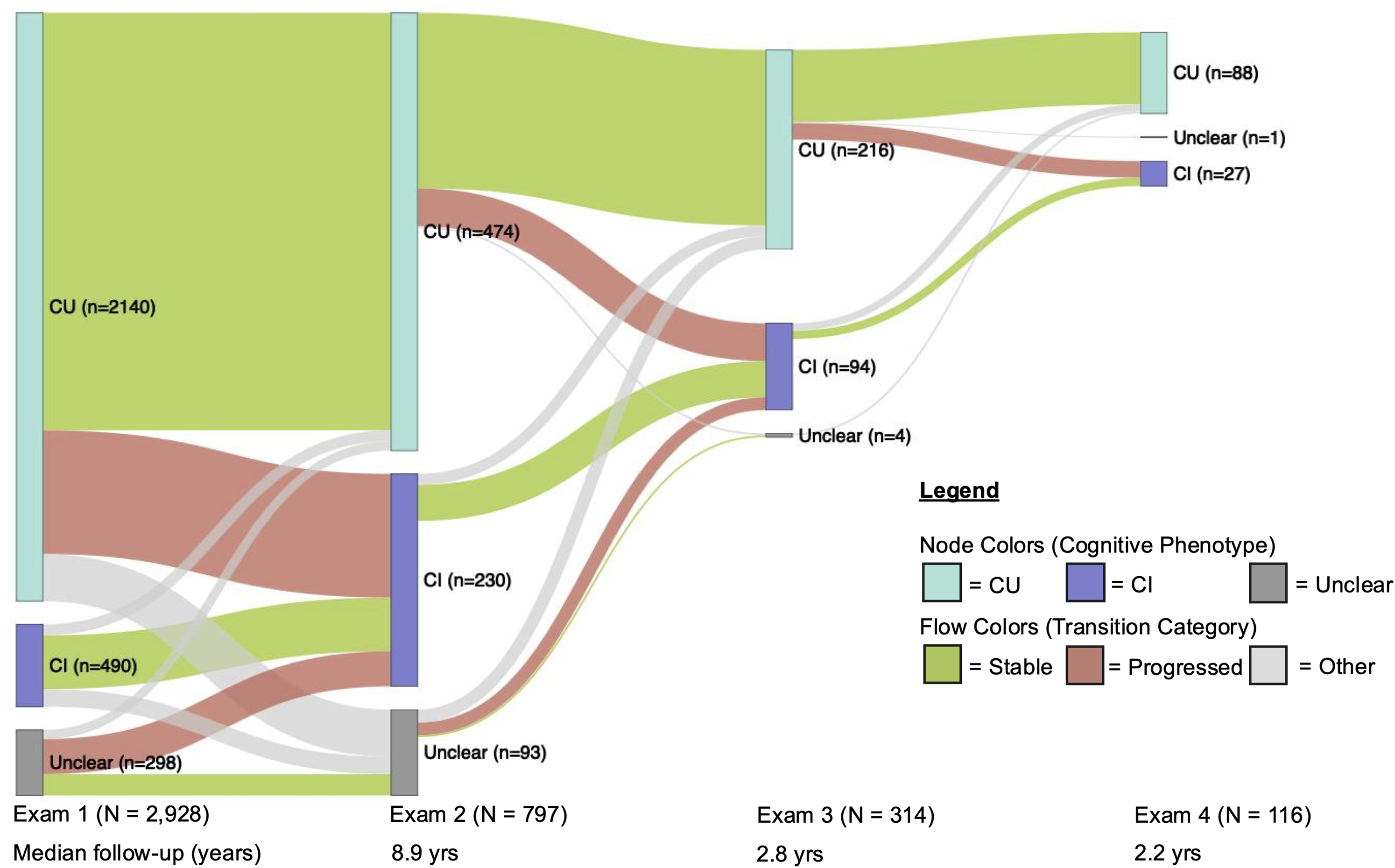
Cognitive trajectories in CAAMP (N = 2,928). The Sankey diagram shows cognitive trajectories of CAAMP participants with at least one cognitive assessment (N = 2,928) across up to four assessments. Cognitive phenotypes were categorized as cognitively unimpaired (CU), cognitively impaired (CI; including Mild Cognitive Impairment [MCI], Cognitively Impaired, not Alzheimer Disease [CINAD] and Alzheimer Disease [AD]), or Unclear. Node size (vertical rectangles) represents the number of participants in each phenotype group and ribbon width (horizontal flows) represents the number transitioning between groups at consecutive examinations. Transitions were classified as: stable (no change in phenotype), progressed (CU/Unclear to CI), or other (all remaining phenotype changes). Exam columns are sequential and not scaled to elapsed follow-up time. Median follow-up was 8.9 years between Exam 1 to 2 (N = 797), 2.8 years between Exam 2 to 3 (N = 314) and 2.2 years between Exam 3 to 4 (N = 116).

CAAMP analyses commonly compare CU controls with both AD cases and the broader CI phenotype. Using these comparisons, among participants with available *APOE* genotype, and AD and CU status, age of onset differed by *APOE* genotype (*ANOVA; p = 0.001*), with *APOE* ε4 carriers exhibiting a younger mean age at onset of 77.8 years compared to non *APOE* ε4 carriers with 80.9 years (Table 3). Findings for CI cases were similar (details in Supplemental Material 3). Together, these findings suggest our observed onset ages tend toward the older end of the reported range in other populations and are consistent with patterns seen in other Non-Hispanic White populations.^38^

**Table 3.**
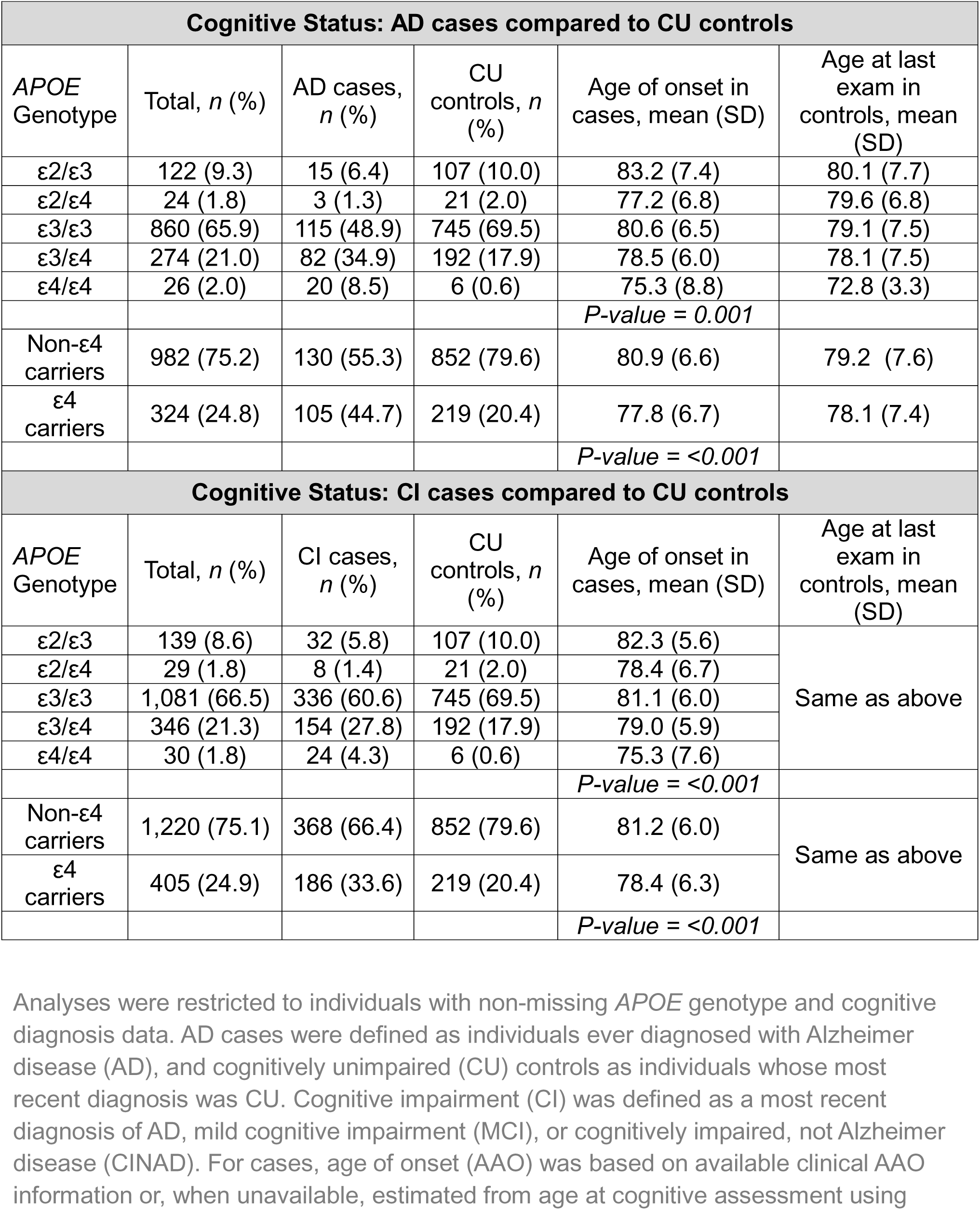
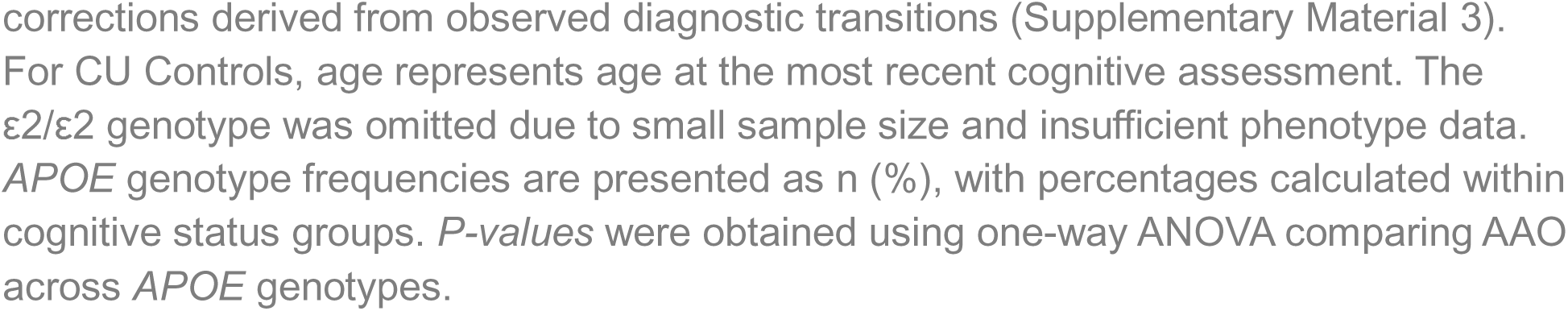
*APOE* genotype distribution by cognitive status, including age of onset and age at last examination.

### 3.2 Amish Pedigrees

For participants new to the study with no enrolled immediate family members, a 3-generation family structure, encompassing all siblings, both parents and all four grandparents, is obtained through the Swiss Anabaptist Genealogical Association (SAGA)^10^, confirmed with the participant at enrollment, and updated at follow-up visits as needed. All 3-generation pedigrees are databased for all participants using Progeny (Progeny Genetics LLC). Given the complexity of connecting all 3-generation pedigrees back to common ancestors over 14 generations and across the 3,978 participants in CAAMP, an all-connecting path (ACP) pedigree is generated using unique Anabaptist Genealogy Database (AGDB)^9^ identifiers, linking all participants across studies into a single pedigree structure of over 11,000 individuals (Figure 3). Pedigree composition and relative pair count across degree of relatedness is summarized in Table 4.

**Figure 3.**
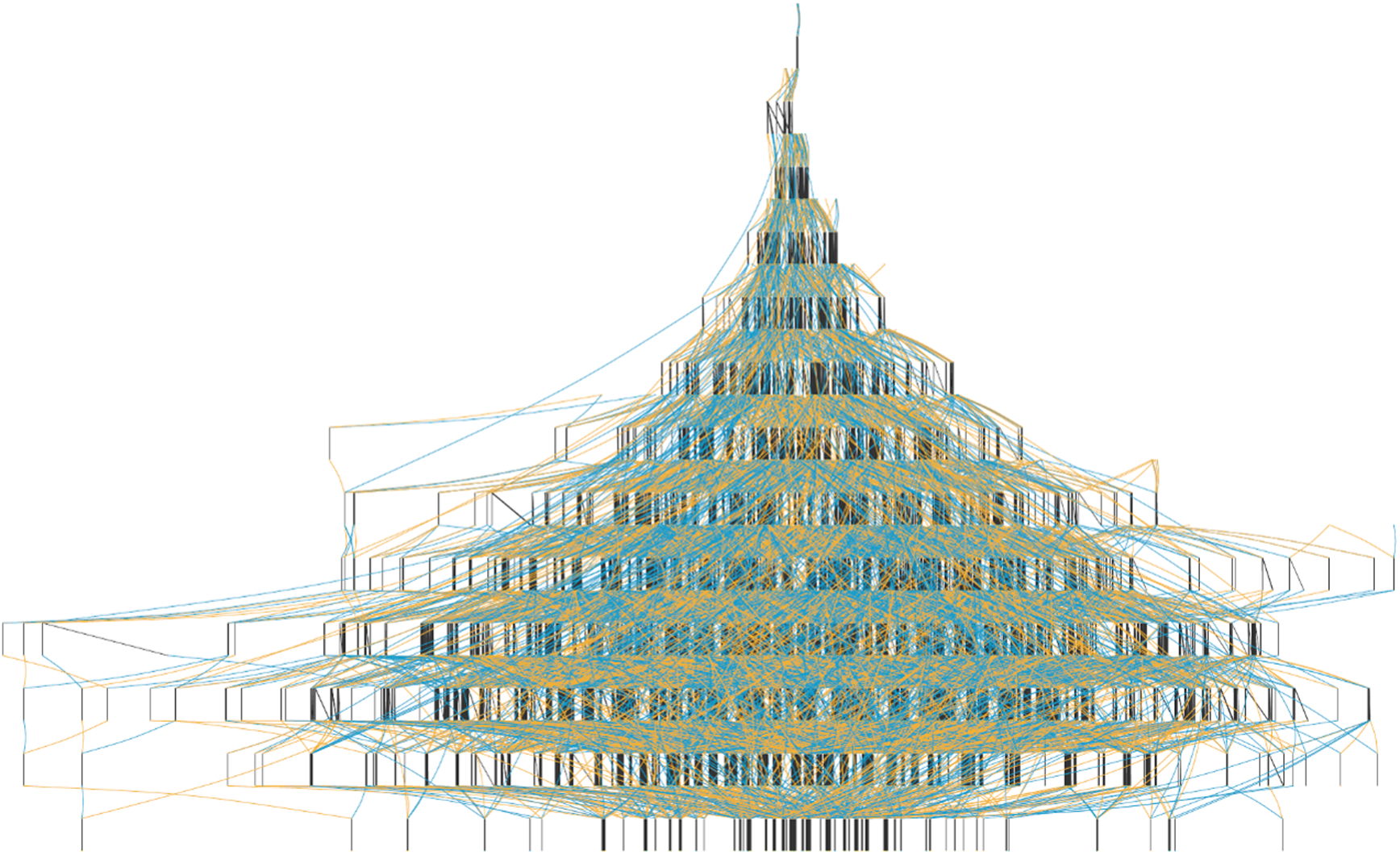
All-Connecting Path Pedigree. Over 11,000 individuals spanning 14 generations are present in this large pedigree, which includes the 3,978 participants in CAAMP. Genealogy data was obtained from the Anabaptist Genealogy Database (AGDB). Colored lines are used to connect generations between families.

**Table 4.**
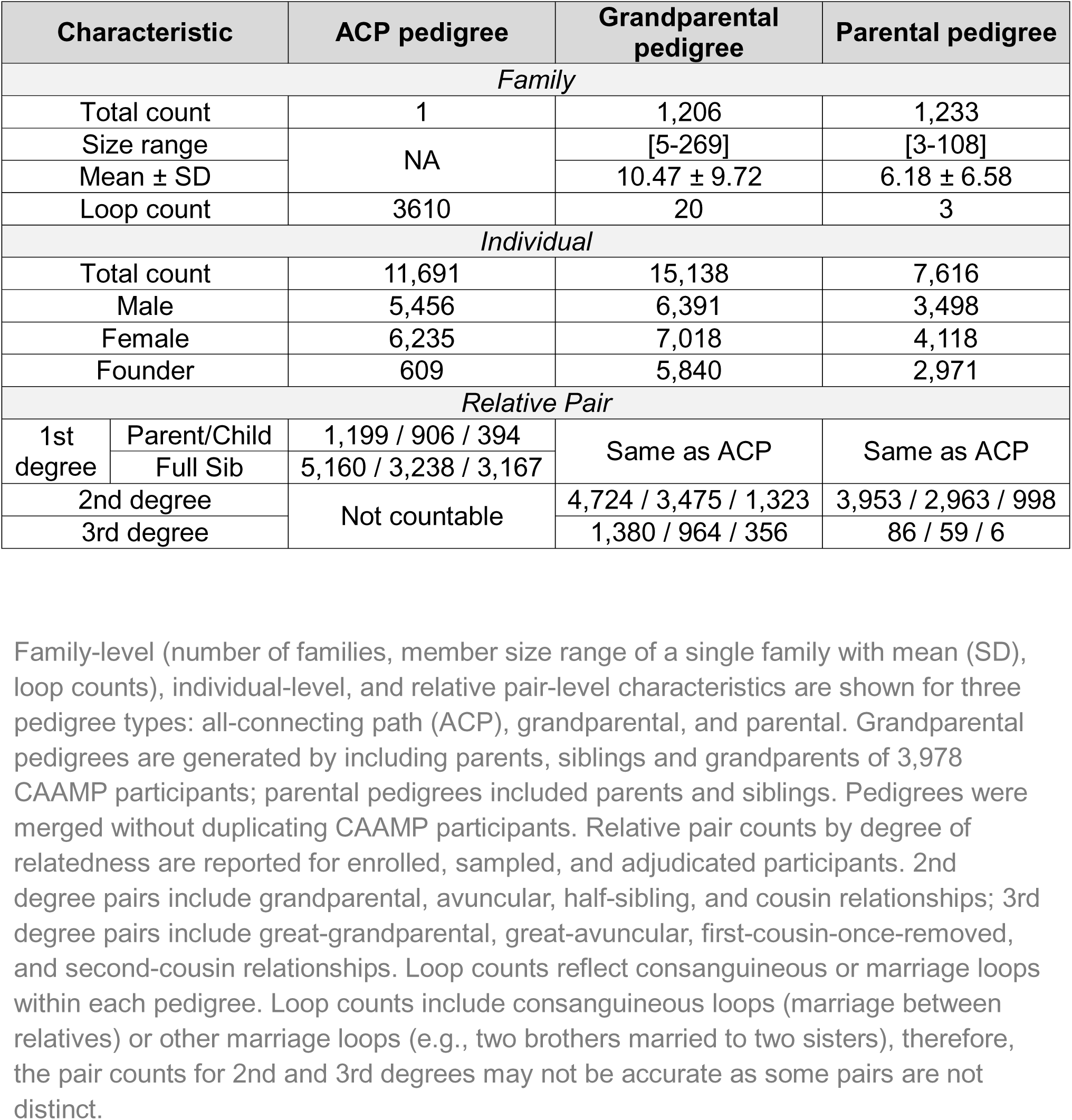
Pedigree characteristics for CAAMP participants (N = 3,978)

### 3.3 Genetic and Biological Resources

Biospecimen samples have been collected longitudinally for participants enrolled in CAAMP. 5,974 samples from 2,842 unique participants have been collected across all sample types, primarily whole blood, DNA and plasma (Table 5). Stratified by site, OH has a total of 3,876 samples from 1,436 unique participants and IN has a total of 2,098 samples from 1,406 unique participants.

**Table 5.**
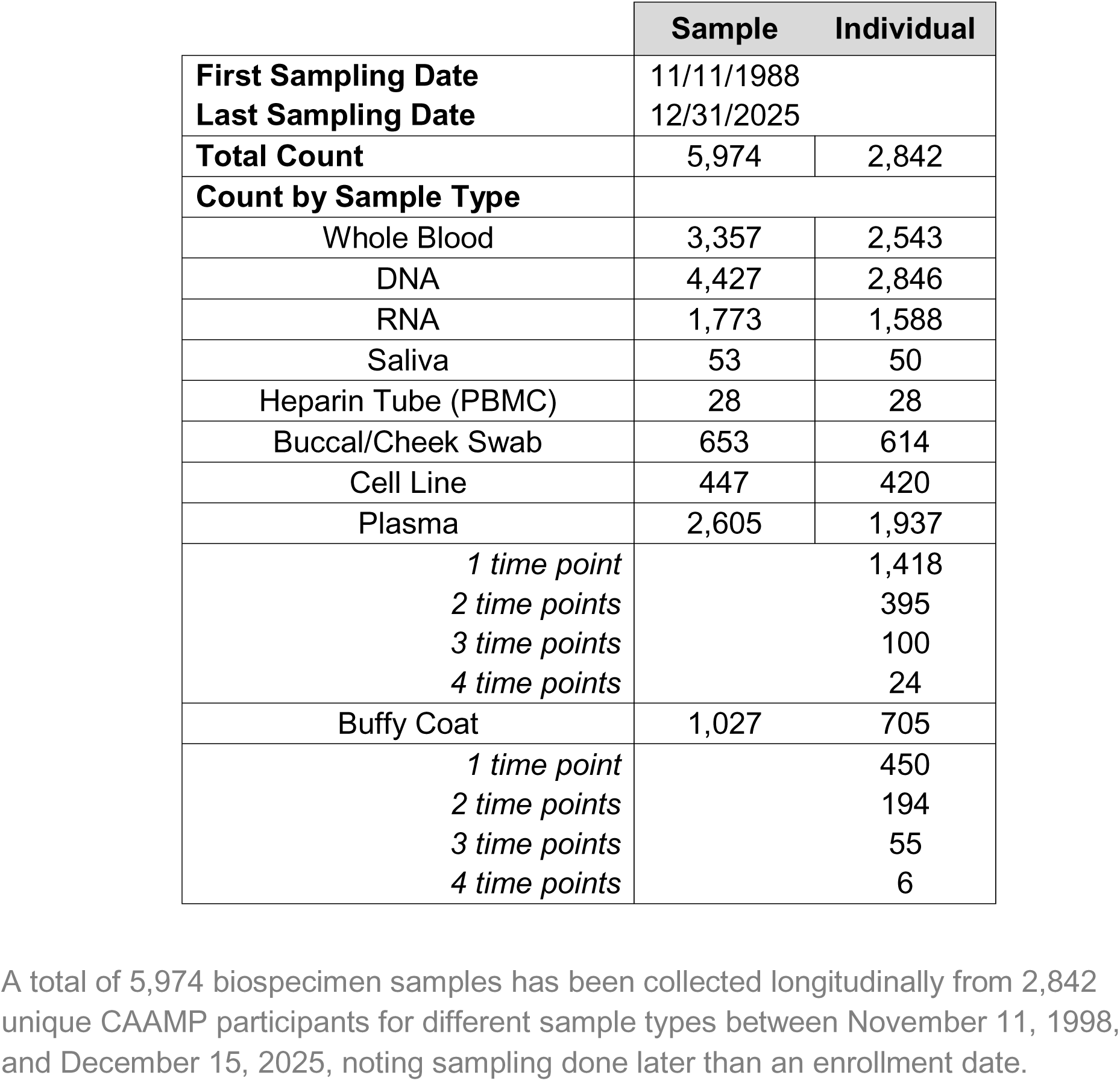
Biological Sample Status for participants by sample type and timepoint.

Plasma and buffy coat samples represent one of the largest longitudinal resources in CAAMP. Plasma samples have been collected longitudinally, with up to four timepoints available for a subset of participants. Buffy coat samples include up to three longitudinal timepoints. The availability of multiple longitudinal timepoints is rare among existing cohorts and enables analyses that are not possible in cross-sectional designs.

Genome-wide genotype data are available for 3,108 samples, generated using either the Illumina Expanded Multi-Ethnic Genotyping Array (MEGAEX) or the Illumina Global Screening Array (GSA); each array has been customized and enriched with variants identified from previous genetic studies done in the Amish or AD. Genotype data underwent standard quality control procedures, followed by imputation using the Michigan Imputation Server^39^ with the Trans-Omics for Precision Medicine (TOPMed) reference panel.^40^ Further details on array customization, quality control thresholds, filtering criteria and imputation procedures are provided in the Supplement Material 4.

Whole genome sequencing (WGS) data are currently available for 1,150 participants using the Illumina NovaSeq 6000 platform. Sequencing alignment for reads was performed against the GRCh38 reference genome, with details on sample and variant level quality control thresholds and filtering criteria provided in Supplement Material 4.

Plasma-based biomarker data is available on 1,138 participants. The biomarkers were quantified using the Quanterix™ HD-X platform (N=808) and the Alamar NULISAseq™ CNS 120 panel (N=330) from 500 μL aliquots stored at -80°C . The Alamar panel includes all Quanterix biomarkers and includes approximately 115 other markers associated with the central nervous system (CNS) (Supplemental Material 4).

### 3.4 CAAMP Contributions to Understanding Aging Related Diseases

While many genetic pathways relevant to aging related diseases are shared with the broader population, studies in the Amish have consistently demonstrated that the relative contribution and interaction of these genetic factors may differ. This makes the Amish population, who are relatively environmentally and genetically homogeneous, particularly valuable to uncovering novel loci and genetic architecture, and identifying mechanisms that may be obscured in more heterogeneous populations.

#### 3.4.1 ADRD

CAAMP has a long history of publications on ADRD, beginning with the 1996 paper demonstrating that the *APOE* ε4 allele was less frequent among Amish cases and controls.^15^ These early findings suggest that while *APOE* is relevant, its role in AD risk may differ between Amish and non-Amish populations. In the mid-2000’s a series of pedigree-based genetic linkage analyses identified both significant and suggestive loci for dementia, specifically on chromosomes 3,7 and 11.^16,41,42^ Together, these studies suggested that the genetic architecture underlying AD risk may be particularly complex and distinct in the Amish.

With the rapid advancement of genotyping chips and sequencing technologies, studies in CAAMP expanded from linkage-based approaches to genome-wide interrogation of genetic variation via genome-wide association studies (GWAS). Cummings et al. 2012 found significant evidence for loci associated with LOAD on chromosomes 2, 3, 9 and 18, with the strongest signal being a SNP in the *CTNNA2* gene on chromosome 2, a gene also implicated in cognitive preservation (see below).^43,44^ Subsequent work by D’Aoust et al. 2015 focused on targeted association testing of exonic variants, with results implicating a *LAMA1* variant in increased LOAD risk, an association specific to the Amish.^45^ More recent studies have shifted toward understanding the broader genetic factors contributing to AD in the Amish. Osterman et al. 2022 demonstrated a smaller contribution of *APOE* to AD risk in the Amish, alongside an overall distinct genetic architecture compared with non-Amish European populations.^17^ Building on this, Osterman et al. 2023 provided early evidence that incorporating population-specific estimates could improve polygenic risk score (PRS) performance for AD when comparing Amish to non-Amish Europeans^46^, highlighting the importance of ancestry-specific modeling in this founder population.

Negative findings include no differences in mitochondrial haplogroups between AD case and control maternal lines, with the results indicating that the etiology of Amish may be distinct from the broader, more outbred European population, suggesting that AD risk may be attributed to nuclear genetic factors, rather than of mitochondrial origin.^47^ More recent negative results indicate no significant correlation of telomere length with cognitive function^48^, *MGMT* was not replicated as a clear risk locus for dementia^49^, and *HFE* and *TF* did not show evidence of a joint association with AD.^50^

Beyond genetics, CAAMP has begun utilizing available blood samples to generate biomarker data. ADRD is a heterogeneous, multifactorial disease with pathophysiological changes often preceding clinical symptoms. This has led to increasing interest and use of plasma-based biomarkers to capture underlying disease processes, though substantial variability exists across individuals. Leveraging pedigree and genotype data, AD-related plasma biomarkers showed moderate heritability, with estimates ranging from 11.1–36.6% using pedigree-based methods and 6.7–28.7% using SNP-based methods.^51^ In CAAMP participants, several plasma biomarkers, including Aβ42/Aβ40, p-tau181, and Aβ42/p-tau181, are potential indicators distinguishing individuals with AD or CI from CU individuals.^52^ Evidence that AD-related plasma biomarkers are moderately heritable and discriminatory in the Amish suggests that this variation is partly genetically driven, supporting the integration of genetic and biomarker analyses to better understand ADRD risk and progression within this cohort.

These findings suggest that AD genetic architecture may be shared across populations, but the importance of specific genetic and biomarker factors may differ between Amish and non-Amish populations.

Beyond genetic or biological factors associated with ADRD and cognitive impairment, CAAMP has also begun studying associations between non-medical drivers of health and cognition, as well as exploring cognitive patterns. Ramos et al. 2021 reported an association between educational attainment and cognitive impairment among the Amish, despite generally lower levels of formal education than in the US non-Amish population, supporting the relationship between education and cognitive outcomes in later life.^53^ Prough et al. 2023 further demonstrated differences between Amish participants and a general European population in aging and ADRD-related changes in verbal and visuospatial memory.^54^ Zaman et al. 2023 used psychometric methods to define more granular cognitive phenotypes in the Amish, offering a more informative alternative to broad clinical classifications and potentially improving power to identify genetic variants associated with cognition.^55^ Taken together, these studies broaden the scope of CAAMP and demonstrate CAAMP’s utility for characterizing cognitive aging in this founder population.

#### 3.4.2 Cognitive Preservation and Successful Aging

An alternative approach toward understanding ADRD is the investigation of protective variants, rather than risk variants. This includes the study of successful aging (SA), a phenotype reflecting the preservation of abilities across multiple domains, cognitive, physical, and functional, at older ages.^56,57^ Concepts such as cognitive resilience and cognitive reserve may also explain heterogeneity in ADRD. With a shift in focus in the late 2000’s and early 2010’s, CAAMP began investigating successful aging in the Amish, identifying linkage to chromosomes 6, 7 and 14^56,57^, and associations with mitochondrial haplogroup X.^58^

In Phase III of CAAMP, ascertainment focuses mostly on enriching for cognitively unimpaired individuals to investigate protective genetic factors in cognition (Figure 1). Several recent studies have identified genetic variants associated with protection from cognitive decline in the Amish, including *SHISA6, CTNNA2*, *LRRTM4*, *WDR12*, and *HIVEP3.*^44,59–61^ Ramos et al. 2023 identified a genome-wide significant association between a *SHISA6* variant on chromosome 17 and delayed cognitive impairment, which was replicated in the NIA-LOAD family-based study and is currently the focus of follow up molecular studies. Main et al. 2024 identified >100 significant linkage signals, including a novel chromosome 2p11.2-13.1 locus associated with cognitive preservation encompassing *CTNNA2* and *LRRTM4*^44^. The lead variant, *rs1402906*, may alter *POU3F2* binding and transcription of these genes, providing a potential mechanism for cognitive preservation.^44^ More recent work by Dorfsman et al.^60,61^ identified additional protective genetic variants in Amish individuals enriched for cognitive preservation despite high genetic risk for AD. These studies identified a linkage signal within *LINC01122* on chromosome 2 and a significant interaction between genetic variants near *KCNA5* and AD genetic risk on chromosome 12, as well as *WDR12*, a previously implicated AD-associated locus on chromosome 2, and *HIVEP3* on chromosome 1 among Amish superagers (aged ≥80 years).^60,61^

This body of work represents a major shift from studying disease risk to understanding resilience, positioning the Amish as a powerful population for identifying factors associated with preserved cognitive function.

#### 3.4.3 Other Age-Related Diseases

CAAMP has also studied Parkinson’s disease (PD), a complex, neurodegenerative disorder of aging, which can share common clinical and pathological features with AD. A genome-wide linkage analysis using microsatellite markers and genotyping known PD genes, examining an eight-generation Amish pedigree, found significant loci (LOD > 3) on chromosomes 3, and 7, with suggestive signals on 10 and 22.^62^ A follow-up study found additional linkage peaks on chromosomes 6, 19, 21 and 22.^63^ Furthermore, genome-wide association and linkage analyses on part of a 4,998 subject pedigree showcased associations in regions near chromosomes 5q31.3 and 10p12.31, with linkage on chromosomes 6 and 10.^64^ In particular, the 5q31.3 region has been identified in previous studies, but this study was the first to find an association of this region with the Amish.

Beyond neurodegenerative diseases, CAAMP has also supported genetic and clinical studies of ocular diseases, particularly AMD and glaucoma. A subset of CAAMP participants has ocular phenotype information on AMD (N = 825) and glaucoma (N = 91). Common variants in the complement factor H (*CFH*) and *ARMS2/HTRA1* loci are among the most extensively studied genetic factors for AMD.^65,66^ In CAAMP participants, a rare *CFH* variant (P503A) detected through exome sequencing showed a significant association with AMD, an association not observed in non-Amish populations.^67^ Subsequent analyses identified additional carriers and suggested that the P503A variant may influence AMD risk through subtle structural effects on the CFH protein, rather than changes in gene or protein expression.^68^ A GWAS of retinal traits and AMD in the Amish identified novel variants significantly associated with drusen measures, including those independent of known AMD risk loci, supporting drusen as a potential biomarker for AMD risk and progression.^69^ Beyond genetic analyses, CAAMP data have enabled collaborative clinical investigations of AMD, including studies of retinal sensitivity, drusen characterization, and structural changes in retinal and choroidal architecture^70–74^, underscoring the translational potential of this cohort.

Similarly, CAAMP studies of glaucoma have leveraged large Amish pedigrees and available comprehensive eye examinations, emphasizing heritable quantitative traits such as vertical cup to disc ratio (VCDR) and intraocular pressure (IOP), which may offer greater power for genetic discovery than disease status alone.^75^

## 4. Discussion

### 4.1 The Legacy of CAAMP

CAAMP’s success is reflected not only in its longevity, but also in the diverse genetic and phenotypic dimensions of successful aging, cognition and other age-related traits characterized in the Midwestern Amish. While sustained collaboration and coordination across institutions have contributed to this longevity, its continued success is fundamentally due to the willingness of participants to participate.

These characteristics have supported development of an extensive biospecimen and data repository within CAAMP. The study has collected multiple biospecimen types, with genetic and biomarker data, including longitudinal plasma samples with up to four timepoints. In parallel, phenotype adjudication has evolved through three phases, improving the depth and consistency of clinical characterization. Together, the scale of the cohort, breadth of biospecimens, genomic biomarker data, longitudinal phenotyping, and multigenerational family structure establish CAAMP as a valuable research resource. This multigenerational family structure can provide additional insights that are not currently possible for other studies about the heritability of risk for the age-related phenotypes being studied by CAAMP. Furthermore, the integration of diverse data types also enables diverse analytical strategies. In addition to internal analyses, CAAMP data have contributed to large collaborative efforts, including the Alzheimer’s Disease Sequencing Project (ADSP).

Important limitations should be considered. Due to cultural considerations and respect for community cultural norms regarding medical procedures and technology, as well as logistical constraints, neuropathological and neuroimaging data are not currently available in CAAMP and rarely conducted in this population. Additionally, analyses must account for familial and genetic relatedness. The sample size is also smaller than that of large biobanks or genetic consortia. However, these same characteristics provide advantages, including the ability to detect rare or moderately frequent DNA variation and to better control for environmental effects. Additionally, the longitudinal depth of CAAMP, with repeated measurements across decades and low attrition, provides a unique sample that cross-sectional biobanks cannot replicate. Notably, while many findings are consistent with those observed in non-Amish European populations, differences in the genetic architecture unique to the Amish have been observed in the phenotypes studied in CAAMP, offering novel, additional insight into disease risk and protective mechanisms.

### 4.2 Future Directions

Continued recruitment and ascertainment will expand longitudinal biomarker data, enabling identification of within-person changes over time. Endophenotypic markers (biomarkers) can provide insight into disease liability and pathophysiological processes, helping to define trajectories and subtypes, predict disease conversion, and estimate age of onset. These efforts may inform therapeutic development and disease management, with repeated measures becoming critical for informing detection in preclinical stages. Given the progressive nature of ADRD and neurodegenerative diseases, longitudinal data are increasingly critical for understanding disease development and progression, and how therapies might be refined to intervene in the upstream, preclinical phase rather than addressing the downstream symptoms once the onset of disease has occurred.

In addition to expanding biological and genomic data collection, CAAMP has begun incorporating measures of non-genetic and non-medical drivers of health to better understand social and environmental influences on disease risk in the Amish, relative to the general population. Social drivers of health are an important and growing area of research, yet their role in ADRD remains incompletely understood. Expanding understanding of how structural and social factors contribute to disparities in ADRD is needed.

Overall, CAAMP provides a valuable resource, opportunity and legacy. Continued study of this founder population offers the potential to identify genetic risk and protective factors while contributing to broader efforts to understand the etiology of ADRD and other age-related diseases, and to inform future therapeutic strategies.

## Data Availability

All data produced in the present study may be available upon reasonable request to the authors, subject to coordination and approval with the respective IRBs.

## Acknowledgements

We are forever grateful to the Amish families for allowing us into their communities and for their continued participation in the research presented and referenced in this manuscript.

We also acknowledge the efforts of all former students, postdocs, staff and faculty who contributed to the CAAMP including Allison Ashley-Koch, Jessica Cooke Bailey, Aneesa Chowdhury, Monique Courtenay, Marilyn Creason, Amy Crunk, Anna Cummings, Laura D’Aoust, EG D’Hondt, Mary Davis, Digna Velez Edwards, Denise Fuzzell, Paul Gallins, PC Gaskell, John Gilbert, Anthony Griswold, Daniel Hahs, Charles Jackson, Lan Jiang, CC Johnson, Stephanie Johnson, Tyler Kinzy, Claire Knebusch, Ioanna Konidari, Charles Kroner, Leighanne Main, Eden Martin, Jacob McCauley, Lynne McFarland, Laura Nations, Michael Osterman, Jairo Ramos, Lori Reinhart-Mercer, JB Rimmler, LC Robinson, Jane Sewell, Y Shao, Susan Slifer, Eric Torstenson, Michael Tramontana, Joelle van der Walt, Andrea Waksmunski, Ping Wang, Weihuan Wang, KA Welsh-Bohmer, Patrice Whitehead, and Andrew Zaman.

In special recognition, we dedicate this work in the memory of Denise Fuzzell and Dr. Charles “Gene” Jackson. Denise’s, dedication to the Amish communities in Ohio and Gene’s dedication to the Amish communities in Indiana were invaluable in fostering the success of CAAMP. Their longstanding dedication to advancing research helped drive our commitment to these communities and our studies.

## Consent Statement

All CAAMP studies were reviewed and approved by the Institutional Review Boards (IRBs) of the institutions responsible for the research at the time the respective study protocols were conducted, and all participants provided informed consent. The current studies are conducted in accordance with the guidelines of the IRBs at Case Western Reserve University and the University of Miami.

