## Supplemental Materials for "A longitudinal study of age-related traits and cognitive function: The Collaborative Amish Aging and Memory Project (CAAMP)"

**Supplemental Material 1: Clinical assessment measures and modifications to neurocognitive assessments to be culturally appropriate for the Amish**

**Clinical assessment measures**

Functional and physical assessments included the Index of Activities of Daily Living (ADL^1^), the Lawton-Brody Instrumental Activities of Daily Living Scale (IADL^2^), the Rosow-Breslau Functional Health Scale^3^, the Nagi Physical Function Scale^4^ and other measures, including gait speed, sit-to-stand and tandem testing^5^.

**Education-adjusted 3MS scores**

In 2021, the ascertainment protocol regarding education was revised. The Modified Mini-Mental State Examination (3MS) scores incorporate possible education-based adjustment. As such, a standardized procedure was developed to assign a single final education value for an individual, if they had multiple exams. Examinations were categorized into two periods: 1) Pre-2021 period, examinations conducted prior to January 1, 2021 and 2) Post-2021 period, examinations conducted on or after January 1, 2021. Participants were then classified into three groups: 1) Education recorded only before 2021, 2) Education recorded only from 2021 onward and 3) Education recorded in both periods. For Group 1, consistent values were retained, but if inconsistencies were present, the most recent recorded education value was assigned. For Group 2, if consistent values were assigned, the recorded education values were assigned, but if there were inconsistencies present, records were manually reviewed. For Group 3, for those who have education values from both periods, the Post-2021 period value was prioritized, reflecting the revised ascertainment protocol. For participants with missing education values, if raw and adjusted 3MS scores were available, the education value was inferred using the 3MS education adjustment scheme (Supplementary Table S1). The difference between the adjusted and raw score was calculated and then mapped to the corresponding education category.

**Supplementary Table 1: Adjustment Scheme for 3MS Using Years of Education**

| Years of Education | Adjustment Points |
| --- | --- |
| <8 | +9 |
| 8-11 | +3 |
| 12 | 0 |
| 13-15 | -1 |
| 16 | -3 |
| >16 | -4 |

**Supplemental Material 2:**

**For Collection of biological samples:**

Like many other aspects of this long-term study, the two data collection sites have developed slightly different ways of approaching the collection of biological samples:

**Ohio:** For the Ohio Amish, sample collection is usually done separate from the collection of phenotypic data. The ascertainment team typically travels together on Wednesdays to subject homes to do the blood collection. This is beneficial for the ascertainment team because they have an extra set of hands for the blood draw and it is efficient because all the samples can be shipped together to CWRU once per week, using overnight delivery service. The study coordinators keep coolers in their vehicles with cool packs to keep the samples cold until they are shipped. Once received in the lab the day after the blood draw, samples are processed and frozen immediately. DNA and plasma aliquots are shipped frozen to the University of Miami at regular intervals. If samples are collected for future generation of iPSCs, these ambient temperature samples are sent directly to UM.

**Indiana:** For the Indiana Amish, sample collection is often completed on the same day as collection of phenotypic data. Prior to 2019, samples were often shipped to the University of Miami the day after collection, so they weren’t processed until 2 days after collection. Beginning in 2019, once biomarker analyses were added to the study protocol, samples were then shipped the same day as collection so that processing could occur the next day. The study coordinators keep coolers in their vehicles with cool packs to keep the samples cold until they are shipped.

**Supplemental Material 3: Age of Onset by APOE Genotype Calculation Details**

A subset of CAAMP participants with phenotype information has age of onset information. All participants are administered a standard battery at each visit consisting of a semi-structured clinical interview and a range of cognitive, behavioral, and functional assessments. In addition, a person who knows the participant well (e.g., spouse, child, or sibling) is interviewed and provides information about current cognitive, behavioral, and functional status and change over time. An important part of the visit is to identify age at which first symptoms, if any, were observed (i.e., age at onset AAO). Both the participant and the informant are asked about changes in memory, function, and behavior and their best estimate of when this change occurred. Clinical coordinators have been trained to query any mention of changes to assess frequency, consistency, and intensity along with corresponding details about potential causal events (e.g., medical conditions). Most important, clinical coordinators query how noticeable the changes are, their impact on daily function, and whether they are part of a decline. All information about estimated age at onset are documented in the research record and are available in the clinical summary provided to examiners.

For those without age of onset information, available transition data of phenotypes was used to calculate a correction value corresponding to a certain phenotype and subtracted from the age at exam. For individuals with transition information about Alzheimer disease cases, the midpoint of 1) the age at the last cognitively unimpaired exam and 2) the age at Alzheimer Disease diagnosis was taken (N=38). The mean correction using transition data on cognitively impaired individuals (N = 190; Alzheimer Disease, Mild Cognitive Impairment and Cognitively Impaired, not Alzheimer Disease) was found to be 4.2 years (SD = 2.9) and 3.4 years (SD = 2.3).

**Supplemental Material 4: Details for Biological and Genetic Data and Quality Control**

**Genotyping**^6^

At the time of enrollment, 30mL of blood is collected from all the participants for use in direct DNA extraction and storage of plasma. Genotype data are collected using an Illumina Expanded Multi-Ethnic Genotyping Array with custom content (MEGAex+3k) or an Illumina Global Screening Array (GSA). The MEGAex chip includes over 2 million markers, whereas the GSA chip included a base quantity of 660,000 markers. When performing chip genotyping, we also included customized content of up to 6,000 variants to the MEGAex chip, including over 1,100 novel variants that were identified from our previous Amish whole-exome sequencing (WES) and whole-genome sequencing (WGS) studies and other associated variants from GWAS and the National Institute on Aging’s Alzheimer Disease Sequencing Project (ADSP) studies that are not already on the chip. After genotype data were attained, imputation was performed based on a Haplotype Reference Consortium (HRC) panel.

Quality control (QC) was performed independently on the MEGAex+3k and GSA genotyping chip sets before merging. For single nucleotide polymorphisms (SNPs), this process included the removal of variants with excess genotype missingness, exclusion of monomorphic and duplicate SNPs, filtering for severe deviations from Hardy-Weinberg equilibrium among common SNPs, and correction of Mendelian errors. For samples, QC involved the removal of individuals based on inconsistent genetic and self-reported sex, as well as an overall genotyping completeness threshold of <5%. Subsequently, imputation was performed using the Michigan Imputation Server and the HRC reference set. The MEGAex+3K and GSA datasets were imputed separately, with submissions using the GRCh38 build for autosomes and hg19 for the X chromosome. The reference population for the HRC was European, and phasing was conducted using the Eagle option. Each dataset underwent a second round of QC after imputation (including filtering by INFO score with a separate threshold for common and rare [minor allele frequency, or MAF, <0.01] SNPs). The two separately imputed sets were then merged using overlapping SNPs.

**Whole Genome Sequencing**^7^

Blood samples are sequenced via the Illumina NovaSeq6000 platform. The WGS was processed following the Alzheimer’s Disease Sequencing Project (ADSP) calling pipelines, and the reads were aligned to GRCh38. The detailed quality control (QC) pipeline used KING 2.3.4 (-autoQC) and PLINK 1.0; samples with sex mismatch, ID mismatch, relationship error, and call rate <99% were excluded. Monomorphic SNPs and SNPs with a call rate <99% were also excluded, and SNPs deviated from Hardy-Weinberg equilibrium threshold of p<1x10-8 were excluded. Only bi-allelic autosomal SNPs and in-dels were included.

**Long Read Sequencing (LRS)**

Blood samples from nine Amish superagers^8^ were selected for long-read sequencing using the PacBio Revio System (Pacific Biosciences). High molecular weight (HMW) genomic DNA was extracted and assessed for quality, including fragment size distribution and DNA damage, using the Femto Pulse System (Agilent Technologies). Based on an internally optimized protocol informed by prior sequencing projects, samples were required to meet the following HMW thresholds for library preparation: ≥70% of DNA fragments >10 kb, ≥50% >30 kb, and ≥10% >42 kb. The pre-sequencing quality control was implemented to maximize HiFi read lengths (kb) and overall HiFi data yield (Gb). Given the small sample size in this experiment, post-sequencing quality control metrics have yet to be fully defined.

DNA quality was assessed using a FemtoPulse system (Agilent Technologies). Samples exhibiting high molecular weight profiles and minimal degradation were selected for library preparation. Genomic DNA was sheared with Megaruptor 3 shearing kit (Diagenode) to target an average fragment size of 15-20kb. Libraries were generated using SMRTbell prep kit 3.0 (Pacbio) per manufacturer’s protocol. Libraries were size selected for material >13 kb with the Lightbench (YourGeneHealth) using a 0.5% agarose ICF size selection cassette and a 10kb Dual dye marker. Final library size was measured with FemtoPulse and concentration was measured by Qubit (Thermofisher Scientific).

The SPRQ Polyermase kit (Pacbio) was used to anneal sequencing primers and bind the sequencing polymerase. One sample was loaded per Pacific Biosciences Revio SMRTcell and each sample was sequenced 1x using a 30-hour movie.

**Quanterix Biomarkers**^9^

Biomarker data were obtained from frozen plasma samples stored in 500 ul aliquots at -80C. Samples were randomly assigned on each plate and run in duplicate. Concentrations of plasma Aβ40, Aβ42, and t-tau were measured using the Simoa Neurology 3-Plex A or Neurology 4-Plex E assays (Quanterix), while p-tau181 was measured using the Simoa P-tau181 Advantage V2 assay. Results for all Aβ and tau biomarkers were above the detection limit of the assays. To ensure the quality of biomarkers used for the analyses, we performed quality control (QC) at the individual biomarker level. We first matched the biomarker samples to clinical data in the Amish. Samples without a consensus diagnosis or from non-Amish individuals were excluded. Some individuals had duplicate results from samples collected on the same date. In such cases, we excluded the set of results with a higher intra-assay coefficient of variation (CV). We then kept the samples with a CV of <20%. Based on summary statistics and visualization, we evaluated samples for each biomarker on a case-by-case basis. The outlier for each biomarker was defined by the median absolute deviation (MAD). This method is more robust in detecting outliers for skewed distributions due to its reliance on non-parametric measures of central tendency and variation.

**ALAMAR Biomarkers**

Plasma from peripheral blood samples from individuals with a corresponding consensus clinical diagnosis (clinical assessment performed within 180 days of the plasma collection date) were analyzed using the NULISA™ 120+ CNS biomarker panel on the ARGO HT System (Alamar Biosciences). The NULISA™ 120+ CNS panel includes a broad set of central nervous system–related protein biomarkers, encompassing all targets previously measured using the Quanterix platform, along with many additional markers. NULISAseq™ multiplex assays leverage next-generation sequencing (NGS) to quantify protein abundance by measuring protein-specific DNA barcodes associated with each analyte. Sequencing reads (FASTQ files) were processed and normalized within the Alamar Control Center analysis software, where they were converted into NULISA Protein Quantification (NPQ) values. NPQ values represent normalized protein measurements derived through multi-step normalization procedures, including correction for assay-specific technical variation using internal controls, plate-level normalization to account for inter-plate variability, and scaling across runs to ensure comparability across the dataset. Internal controls and calibrators were included on each plate for quality assessment, and samples failing predefined quality control criteria were excluded from downstream analyses.

Biomarker-level quality control was performed to identify high-confidence analytes for downstream analysis. Biomarkers were retained if they demonstrated low inter-marker correlation (maximum pairwise correlation coefficient < 0.4), minimum detectability >50% across samples, and mean or median raw read counts >500. Statistical significance was determined using a false discovery rate (FDR) threshold of 0.05 to account for multiple hypothesis testing and to control the expected proportion of false-positive results among significant findings.

**Supplementary Table 2. CAAMP Funding Sources**

| **Dementia/Cognition studies** | **Period** | **Funding Source** |
| --- | --- | --- |
| APOE in Adams County Amish | 1995-1996 | Institutional funds |
| Family Studies of AD in the Ohio Amish | 1999-2000 | Vanderbilt University Discovery Grant |
| Genetic Studies of Dementia in the Amish | 2000-2012 | NIH R01 AG019085 |
| Genetic Study of Successful Aging in the Amish | 2001-2002 | Duke University Pilot grant |
| Genetic Studies of Successful Aging in the Amish | 2003-2007 | NIH R01 AG019726 |
| Protective Genetic Variants for Alzheimer Disease in the Amish | 2017-Present | NIH R01 AG058066 |
| **Other Studies** |  |  |
| Genetic Analysis of Parkinsonism in an Ohio Amish Family | 2003-2004 | NIH /K08 NS044298 |
| Parkinson’s Disease in the Ohio Amish | 2006-2009 | Michael J. Fox Foundation Grant |
| Genetic Examination of AMD in the Midwestern Amish | 2008-2011 | Brightfocus M20080540 |
| Genetic Epidemiology of AMD in the Old Order Amish | 2013-2018 | NIH R01 EY023164 |
| Epidemiology of Biomarkers of AMD Progression | 2022-Present | NIH R01 EY030614 |
